# Utilizing early-prediction of respiratory infections in Larissa, Thessaly, Greece: Incorporating daily school absenteeism as an early-warning indicator

**DOI:** 10.64898/2026.07.12.26357892

**Authors:** Meletis Eleftherios, Rousogianni Eleni, Poulakida Irene, Perlepe Garyfallia, Boutlas Stylianos, Papadamou Georgia, Papagiannis Dimitrios, Konstantinos Kapsalis, Pavle Banovic, Lioupi Olympia, Gourgouliannis Konstantinos, Kostoulas Polychronis

## Abstract

**Background:** The outbreak of COVID-19 in Greece prompted extensive public health measures, including the first national lockdown and the suspension of in-person schooling. Recognizing the significant role of children in community transmission due to their contacts in schools, school absenteeism data began to be systematically recorded as a potential indicator of outbreak patterns.

**Objectives:** This study aims to explore the utility of incorporating school absenteeism data in an early warning surveillance system for respiratory infections, particularly in predicting the onset and spread of diseases such as COVID-19 and influenza.

**Methods:** We utilized school absenteeism data from primary schools and kindergartens in the Municipality of Larissa for the 2022–2024 school years, alongside health data from the University Hospital of Larissa (UHL). These included incidence rates of respiratory infections, COVID-19, and flu cases, which were cross-referenced with absenteeism patterns.

**Results:** The analysis showed that peaks in absenteeism often preceded increases in cases of respiratory infections, COVID-19, and flu, suggesting absenteeism as a potential early warning indicator. Notable divergences in patterns were observed during school closures for holidays, which posed challenges in data continuity and surveillance effectiveness.

**Conclusions:** School absenteeism data significantly enhances the capability for early detection and monitoring of respiratory disease outbreaks. To improve future surveillance and outbreak prediction, integrating more comprehensive data sources and refining predictive models to accommodate educational calendar variations is recommended.

## Introduction

The first case of COVID-19 in Greece was reported on 26 February 2020 (Kousi et al., 2021). Soon afterward, in March 2020 public health officials imposed the first national lockdown, which included the suspension of in-person schooling and transition to online lessons. At that time, children were considered less susceptible to infection, typically experiencing only mild symptoms. However, they were recognized as playing a significant role in community transmission, due to their numerous contacts in settings such as childcare centers and schools (Politis et al., 2021). Following the first lockdown in the spring of 2020, Greek officials introduced a four-level preparedness map with corresponding colors designed to communicate the risk levels to residents and visitors and to guide the associated public health measures. At the two highest risk levels, public health measures, such as alternating the opening hours of catering and entertainment centers and suspending schools, were implemented at the regional or municipal level (COVID-19 in Greece). Consequently, schools began to systematically record and report the daily number of student absences as an indicator of potential outbreaks (World Health Organization, 2020).

Moreover, several, simple and more sophisticated, early-warning methods were developed to predict upcoming epidemic waves, based on the number of daily COVID-19 cases. For instance, the trend of the weekly number of new COVID-19 hospitalizations per 100,000 inhabitants is a gross indicator. Furthermore, Kostoulas et al., 2021, designed a statistical early-warning tool called Epidemic Volatility Index (EVI), originally implemented for predicting COVID-19 epidemic wave. Since then, EVI has been applied in other settings for predicting epidemic waves. Meletis et al., 2024 describes a pilot application of EVI to data from the pulmonary clinic of the university hospital of Thessaly, Greece for monitoring and wave-prediction for respiratory infections, COVID-19, and flu cases.

The presented work aims to explore school absenteeism data as a granular view of health trends. Children frequently suffer from gastrointestinal and respiratory illnesses, leading to school absences. As key transmitters of infections, they spread illness in schools, which then extends to the wider community through close contact with family members like parents and grandparents (Donaldson et al., 2021). An uptick in school absences can be an early, actionable alert for public health officials and school administrators, especially as children are often among the first to be affected by seasonal and pandemic illnesses, with absences starting from the very first day of symptoms (Donaldson et al., 2021). This can provide valuable warning before clinical cases are confirmed or widespread community transmission is detected. This lead time is crucial for implementing measures like increased hygiene practices, temporary school closures, or targeted testing. Analytically, we examined the trend of daily school absenteeism records and evaluated against the output of EVI to data on respiratory infections collected from the from the pulmonary clinic of university hospital of Thessaly (UHL).

## Methods

School absenteeism data from all the primary schools and kindergarten in the Municipality of Larissa for school years 2022 – 2023 and 2023 up to 5 April 2024 were provided from the Regional Directorate of Primary & Secondary Education of Thessaly. To safeguard individual privacy and adhere to ethical guidelines, anonymized and aggregated data on the total number of absences per school per day were collected—no personal identifiers were shared. According to duty schedule, UHL receives patients in 24h-turns (i.e., on every second day) and therefore the number of respiratory infections, COVID-19 and flu cases was available on every two days. Among patients admitted in the Emergency Department of university hospital of Thessaly, Greece, data are collected for those who meet at least one of the following inclusion criteria: (i) cough (productive or not), (ii) malaise and fatigue (iii) headache (iv) body temperature above 37.3 °C (v) sneezing (vi) congested nose (vii) dyspnea/shortness of breath (viii) chest tightness. Exclusion criteria of the study include:(i) known infection at another site that explains symptoms (ii) inability to cooperate (iii) denial of participation. Polymerase Chain Reaction (PCR) or antigen-specific tests for SARS-COV2 and flu virus as well as chest X-ray are performed in all participants that fulfill the inclusion criteria. Any positive test (PCR or antigen-based test) is recorded as COVID-19 or flu infection respectively. Participants with negative tests and relative respiratory symptoms are considered as cases of respiratory infections in general. Patient data used in this study were anonymized (i.e., personal identifiers were removed), ensuring that individual patient information remained confidential and in compliance with regional and international data protection regulations.

The 14-day moving average of the sum of absences from primary schools and kindergarten was considered as the early-warning indicator. In this study the trend of the 14-day moving average of absences was evaluated against the output from the ongoing pilot application of EVI to data from the pulmonary clinic of UHL. The 14-day moving average was employed to smooth out daily fluctuations in absence data, notably accounting for weekends when schools are not in session, and to maintain consistency with methodologies used in the ongoing pilot application of EVI at the UHL.

## Results

The output of EVI from the data on respiratory infections, COVID-19, and flu cases, collected from the from the pulmonary clinic of UHL is reported in an online application. Figures 1, 2, 3 display the combined output of EVI’s application to the UHL and the trend of the school absences. Red dots correspond to dates that, according to EVI, an early warning was issued, while grey dots correspond to time points without an early warning indication. There is no distinct difference of the absenteeism patterns between primary schools and kindergarten, except from the period between December 2022 and January 2023. During this time, the number of absences for the primary schools increased, while it decreased for kindergartens.

**Figure 1.**
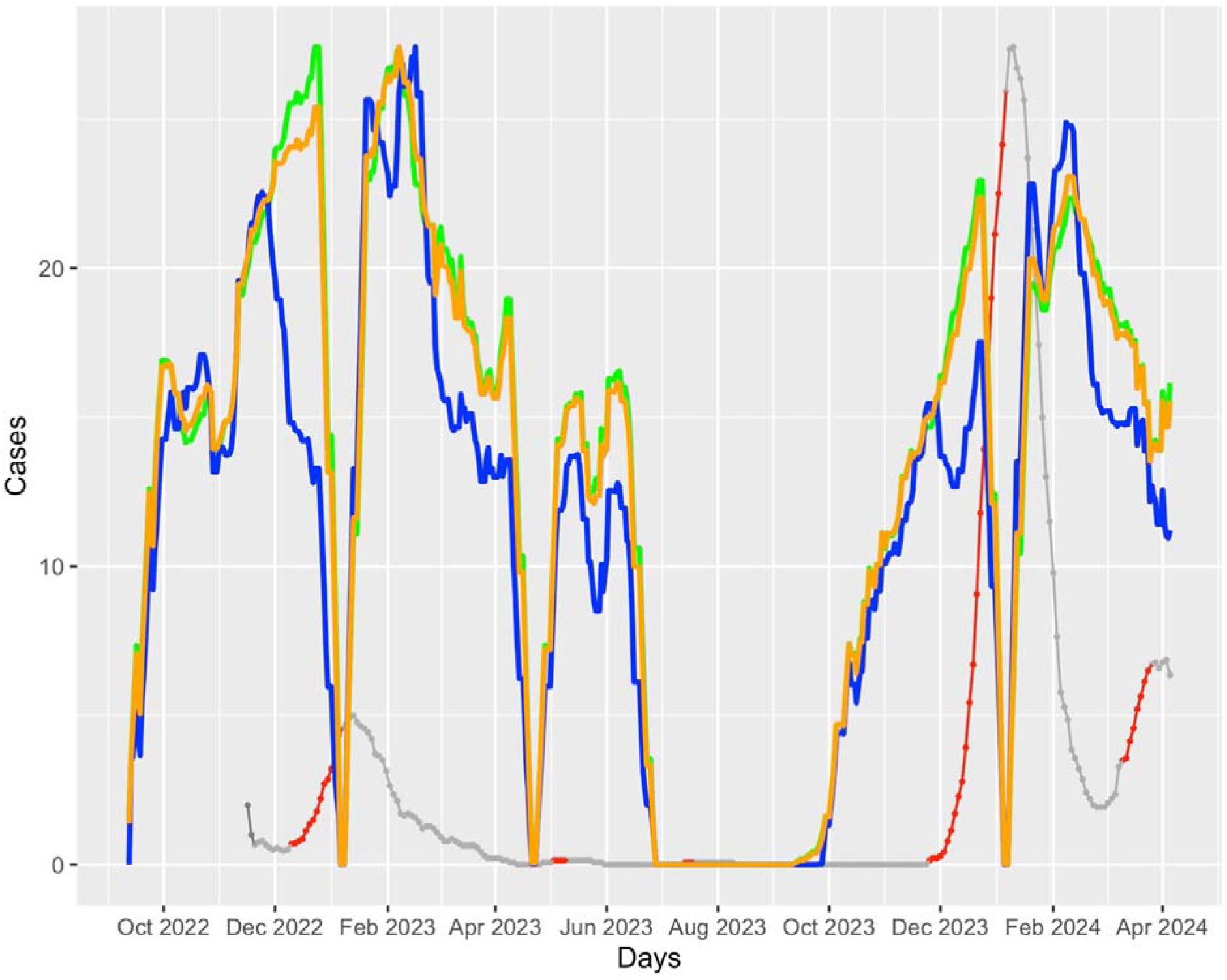
Trends in school Absenteeism and EVI Output on Respiratory Infections from October 2022 to April 2024. Green line represents absenteeism trends in primary schools, blue line in kindergartens, orange line the combined absenteeism trends of primary schools and kindergartens. Red dots refer to time points when EVI issued an early warning signal, while grey dots to point when it did not issue an early warning signal. The drop to zero in school absenteeism in January 2023, late April 2023, and June-August 2023 corresponds to Christmas, Easter, and summer holidays, respectively.

**Figure 2.**
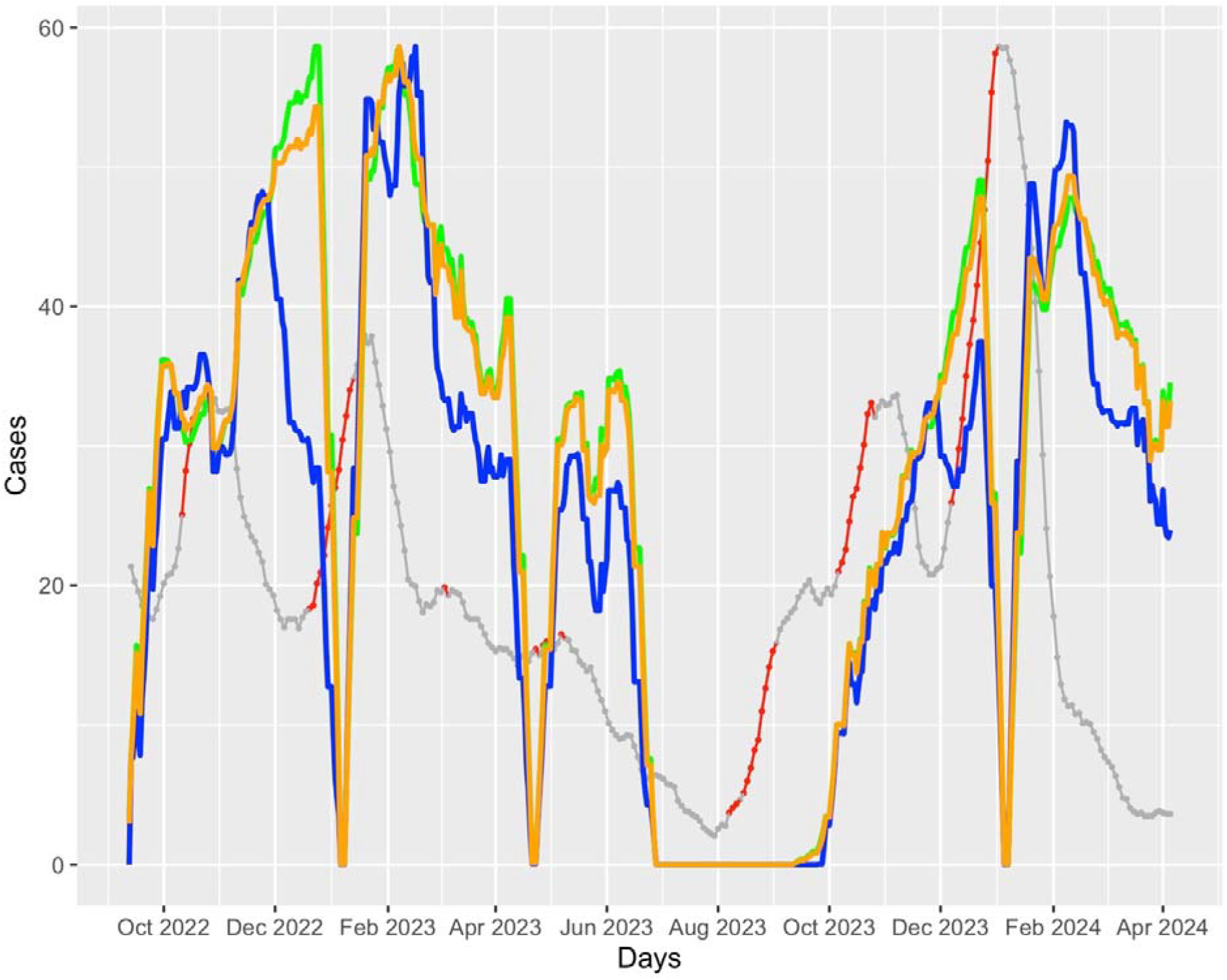
Trends in school Absenteeism and EVI Output on COVID-19 cases from October 2022 to April 2024. The drop to zero in school absenteeism in January 2023, late April 2023, and June-August 2023 corresponds to Christmas, Easter, and summer holidays, respectively.

**Figure 3.**
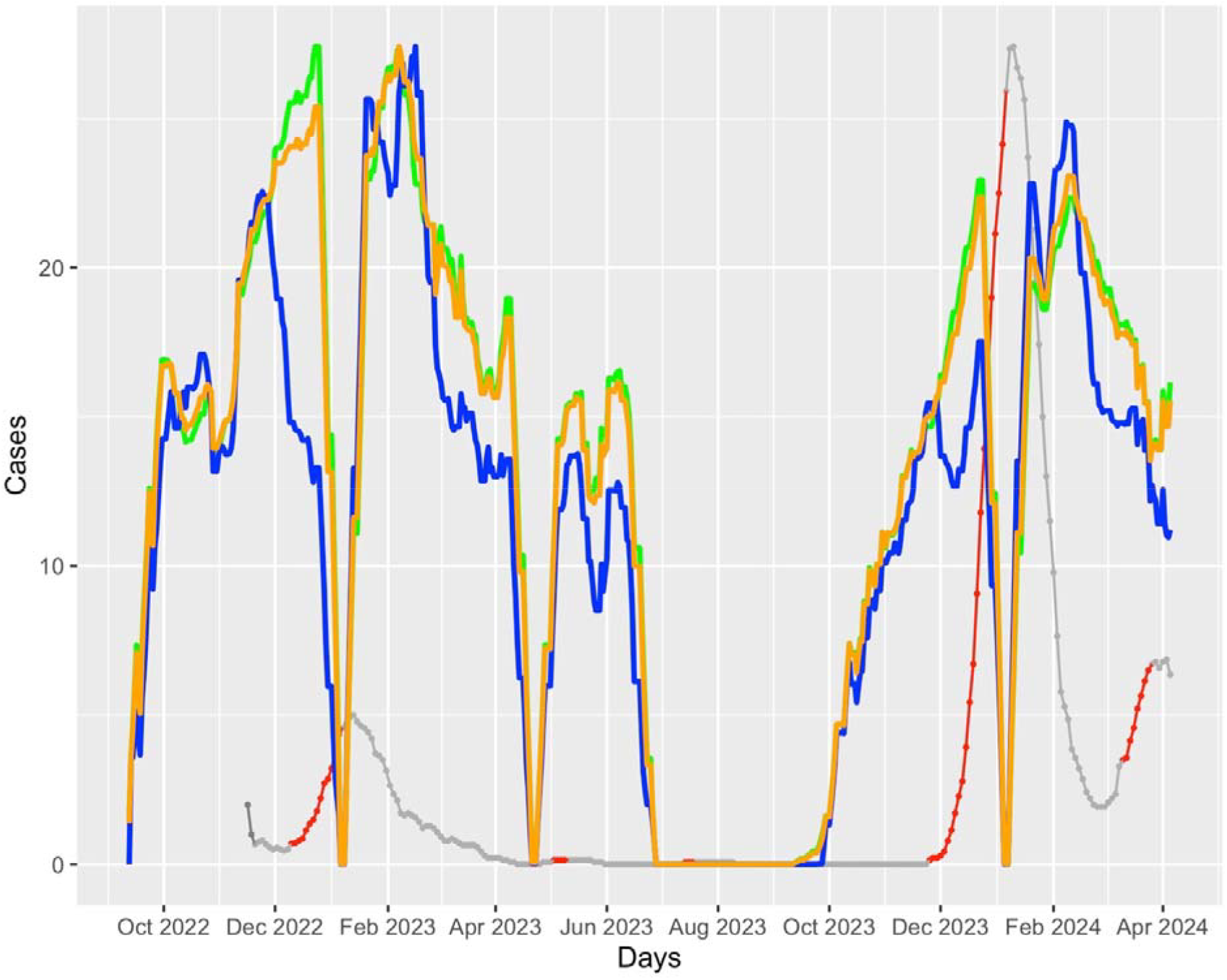
Trends in school Absenteeism and EVI Output on flu cases from October 2022 to April 2024 The drop to zero in school absenteeism in January 2023, late April 2023, and June-August 2023 corresponds to Christmas, Easter, and summer holidays, respectively. *Absenteeism data for primary schools, kindergartens, and their combined total were scaled to the maximum number of cases in each figure to allow direct comparability on the graph.

Inclusion of daily school absenteeism data allows for more precise monitoring and trend analysis for respiratory infections and beyond. Notably, the increasing trends in absenteeism corresponded the periods when early warnings were issued from the online application implemented to the pulmonary clinic of UHL. More specifically, peaks in absenteeism often preceded increases in respiratory infections (November 2022, October 2023 – January 2024), COVID-19 cases (October 2022, November – December 2023) and particularly flu cases (December 2022, October 2023 – January 2024). These results promise a crucial lead time for healthcare preparedness, aiding in targeted public health interventions. On the other hand, there are periods, such as December 2022 (Christmas holidays) and August 2023 (summer holidays), where the patterns diverge due to school closures; consequently, absenteeism data are not recorded during these times.

## Discussion

Our study demonstrates that the integration of school absenteeism data into public health surveillance significantly enhances the early detection of respiratory infections. The correlation between increased absenteeism and subsequent rises in infection rates underscores the potential of school absenteeism as a real-time, proactive surveillance tool. This finding is particularly relevant in the context of the ongoing challenges posed by seasonal and pandemic respiratory diseases, as early detection is critical for timely public health interventions and resource allocation.

The predictive framework has shown valuable potential use in forecasting spikes in respiratory infections. This accuracy is evident from the alignment of increased absenteeism with peaks in reported cases, validating the efficacy of EVI for broader respiratory surveillance. This suggests that integrating similar predictive tools in routine public health strategies could be beneficial, especially in regions where digital health infrastructures can support real-time data analysis. From a bioethical perspective, early detection leads to proactive action. In line with the principle of beneficence it enables interventions—such as improved hygiene protocols, targeted testing, or temporary school closures—that can protect vulnerable populations and help contain outbreaks (Kostkova, 2018). At the same time, public health strategies must remain transparent about how data are collected, processed, and utilized in order to preserve trust and legitimacy in the community. Moreover, medical mediation between relevant stakeholders can provide the framework for the design of more efficient control programs because such programs both address the needs of the community and uphol consensus (Richard et al., 2022).

However, the predictive accuracy of absenteeism data for flu cases presented challenges, particularly due to the pronounced seasonality of flu outbreaks (Jones, 2020). Unlike more uniformly occurring respiratory infections, flu cases peak sharply during colder months, and their predictability may be obscured by holiday-related school closures as observed in December 2022 and 2023. The divergences in absenteeism patterns during school closures such as the Christmas and summer holidays also pose interpretative challenges (Jackson et al., 2014). These closures disrupt the continuity of absenteeism data, highlighting a limitation in the utility of school-based data. These periods require alternative surveillance strategies as traditional school-based monitoring is unavailable. Future frameworks could benefit from incorporating more diverse data sources during these periods to maintain surveillance continuity.

Given the effectiveness of the absenteeism-based surveillance illustrated in our study, there is a compelling case for broader implementation of this approach. Expanding the framework to include other educational institutions and perhaps even workplaces could provide a more comprehensive view of community health trends. Additionally, further research should explore the integration of environmental and socio-economic data to refine the predictive accuracy during typically data-sparse periods like school holidays (e.g., Copernicus Relay Network in Greece).

A limitation of this study is that we assessed all-cause school absenteeism rather than influenza-like illness (ILI)-specific absenteeism, which has a stronger correlation with community surveillance of respiratory infections (Tsang et al., 2023). While ILI-specific measures could improve accuracy, they require additional resources for tracking the reasons for absences, which may delay data collection and reduce school participation (Tsang et al., 2023). Additionally, absenteeism can be influenced by non-infectious factors, but this is less relevant to our study since younger children, the focus of our research, are more likely to miss school due to illness compared to older children, who often miss school for other reasons. Another potential limitation is the possibility of biases in absentee reporting and, the reliance on symptomatic reporting in schools may underrepresent asymptomatic cases, particularly in diseases like COVID-19.

## Conclusion

In conclusion, while school absenteeism data significantly enhances the surveillance and prediction of respiratory infections, the approach requires adjustments to address seasonal variations and data gaps due to school closures. Future efforts should focus on integrating additional data layers and exploring advanced analytical models to overcome these challenges.

## Data Availability

The data presented in this study are available on request from the corresponding author.

http://83.212.175.101:3838/version10/

